# Event Rate and Predictors of Post-Acute COVID-19 Sequalae and the Average Time to Diagnosis in General Population

**DOI:** 10.1101/2023.02.23.23286336

**Authors:** K. John Muthuka, M. Caleb Mutua, M. Japheth Nzioki, Rosemary Nabaweesi, J. Kelly Oluoch, K. Michael Kiptoo

**Affiliations:** Kenya Medical Training College, Nairobi, Kenya; Jumeira University, Dubai, United Arabs Emirates; Kenya Medical Research Institute, Nairobi, Kenya; Meharry Medical College, NT, United States; South Eastern Kenya University, Kitui, Kenya

## Abstract

**Background:** Post- COVID-19 sequalae involves a variety of new, returning or ongoing symptoms that people *experience more than four weeks after* getting COVID-19. The aims of this meta-analysis were to assess the prevalence of Post-Acute COVID-19 sequalae and estimate the average time to its diagnosis; and meta-regress for possible moderators.

**Methods:** A standard search strategy was used in PubMed, and then later modified according to each specific database. Search terms included “long COVID-19 or post-acute COVID-19 syndrome/sequalae”. The criteria for inclusion were published clinical articles reporting the long COVID-19, further, the average time to diagnosis of post-acute COVID-19 sequelae among primary infected patients with COVID-19. Random-effects model was used. Rank Correlation and Egger’s tests were used to ascertain publication bias. Sub-group, sensitivity and meta-regression analysis were conducted. A 95% confidence intervals were presented and a p-value < 0.05 was considered statistically significant. Review Manager 5.4 and comprehensive meta-analysis version 4 (CMA V4) were used for the analysis. The trial was PROSPERO registered (CRD42022328509).

**Results:** Prevalence of post-acute COVID-19 sequalae was 42.5% (95% confidence interval (CI) 36 % to 49.3%). The PACS event rates’ range was 25 % at four months and 66 % at two months and mostly, signs and symptoms of PASC were experienced at three (54.3%, P < 0.0001) to six months (57%, P < 0.0001), further increasing at 12 months (57.9%, P= 0.0148). At an average of two months, however with the highest event rate (66%), it was not significantly associated with PACS diagnosis (P=0.08). On meta-regression, comorbidities collectively contributed to 14% of PACS with a non-significant correlation (Q = 7.05, df = 8, p = 0.5313) (R^2^=0.14). A cardiovascular disorder especially hypertension as a stand-alone, showed an event rate of 32% and significantly associated with PACS, 0.322 (95% CI 0.166, 0.532) (P < 0.001). Chronic obstructive pulmonary disorder (COPD) and abnormal basal metabolic index (BMI) had higher event rates of PACS (59.8 % and 55.9 %) respectively, with a non-significant correlation (P > 0.05). With a significant association, hospital re-admission contributed to 17% (Q = 8.70, df = 1, p = 0.0032) (R^2^= 0.17) and the study design 26% (Q = 14.32, df = 3, p = 0.0025) (R^2^=0.26). All the covariates explained at least some of the variance in effect size on PACS at 53% (Q = 38.81, df = 19, p = 0.0047) (R^2^ analog = 0.53).

**Conclusion:** The prevalence of PACS in general population was 42.5%, of which cardiovascular disorders were highly linked with it with COPD and abnormal BMI also being possible conditions found in patients with PACS. Hospital re-admission predicted highly, an experience of PACS as well as prospective study design. Clinical and methodological characteristics in a specific study contributed to over 50% of PACS events. The PACS event rates ranged between 25 % at four months and 66 % at two months with most signs and symptoms experienced between three to six months increasing at 12 months.

## Introduction

Post-acute sequelae of COVID-19, also known as “long COVID,” is used to describe the long-term symptoms that might be experienced weeks to months after primary infection with SARS-CoV-2, the virus that causes COVID-19 ^1^. Recent literature suggests that, the syndrome is described by a diverse set of symptoms that persist after a diagnosed COVID-19 infection^2^. This post-acute infection represents a significant challenge for patients, physicians, and society because the causes, patient profile, and even symptom patterns remain difficult to characterize^3^. It may include memory loss, gastrointestinal (GI) distress, fatigue, anosmia, shortness of breath, and other symptoms. PASC has been associated with acute disease severity^4^, further, it is suspected to be related to autoimmune factors^5^, as well as unresolved viral fragments^6^. Post-COVID conditions are found more often in people who had severe COVID-19 illness, but anyone who has been infected with the virus that causes COVID-19 can experience post-COVID conditions, even people who had mild illness or no symptoms from COVID-19^7^. There is no test to diagnose post-COVID conditions, and people may have a wide variety of symptoms that could come from other health problems. This can make it difficult for healthcare providers to recognize post-COVID conditions. Diagnosis is considered based on health history, including if you had a diagnosis of COVID-19 either by a positive test or by symptoms or exposure, as well as doing a health examination^8–10^. Long COVID may be due to persistent immune disturbances^11^.

Studies of patients who have recovered from SARS-CoV-2 infection but have persistent symptoms have ranged widely in size, quality, and methodology, leading to confusion about the prevalence and types of persistent symptoms^12^. SARS-CoV-2 can produce short/long-term sequelae and reports describing post-acute COVID-19 syndrome (PACS) in the general population are increasing, however limited by lack of proper pooled average time estimation for diagnosis or occurrence. Therefore, this review aimed at assessing the prevalence and factors associated with PACS in our cohort of general population.

## Methods

A standard search strategy was used in PubMed, and then later modified according to each specific database to get the best relevant results. These included MEDLINE indexed journals; PubMed Central; NCBI Bookshelf and publishers’ Web sites. The language was restricted to English and the search dates were for studies conducted/published between June 2021 and August 2022. The basic search strategy was built based on the research question formulation (i.e., PICO or PICOS)^13^. They were constructed to include free-text terms (e.g., in the title and abstract) and any appropriate subject indexing (e.g., MeSH) expected to retrieve eligible studies, with the help of an expert in the review topic field or an information specialist. The summary of search terms was; long COVID-19; long COVID-19 syndrome; post-Corona Virus syndrome; Post-acute COVID-19 sequelae; Post-acute COVID-19 syndrome; Post-acute COVID-19 condition. Because the study for this topic would be limited, the outcome term (long COVID-19) in the search term was not be included initially in order to capture more studies. Preferred Reporting Items for Systematic Review and Meta-Analysis (PRISMA) statement.^14^ was used. All identified article titles and abstracts were screened independently by two authors (JKM, JMN), with those meeting the inclusion criteria screened further by full text review. On occasions when it was not clear from the abstract if studies were of relevance, the full text of the article was reviewed. Unanimous consensus was met on the inclusion of proposed studies for full text review among the authors (JKM, JMN, KO and CM). Full text studies were further evaluated against the inclusion and exclusion criteria. The reference lists of included studies were reviewed to ensure no other relevant studies were overlooked.

### Search terms and criteria for inclusion

Search terms included for PubMed was as follows: (long COVID-19; long COVID-19 syndrome; post-Corona Virus syndrome; Post-acute COVID-19 sequelae; Post-acute COVID-19 syndrome; Post-acute COVID-19 condition) AND (“the study” [Publication Type] OR “study as the topic” [MeSH Terms] OR “study” AND [All Fields]). A search limit for articles published from mid-2020 applied. The criteria for inclusion were published research articles reporting: 1) the rate prevalence of post-acute COVID-19 syndrome (at admission or following admission) in general population 2) the possible associated clinical parameters. Studies were excluded if they were case reports, review articles, conference abstracts, non-clinical studies, or were not available in the English language.

### Data extraction

The included studies were evaluated for the authors, year of publication/ the study was conducted, title, where it was conducted, study design (prospective, retrospective or other), age and sex of patients, number of patients, number of post-acute COVID-19 sequalae and the time of diagnosis of post-acute COVID-19 sequalae (on admission or following admission with mean/ average time to diagnosis in months). A post-acute COVID-19 sequalae case was defined as the illness that occurs in people who have a history of probable or confirmed SARS-CoV-2 infection; usually within three months from the onset of COVID-19, with symptoms and effects that last for at least two months.

### Outcome measures

The primary objectives were to report the prevalence of post-acute COVID-19 sequalae using the most recent data, post-acute COVID-19 sequalae in PLWH as from mid-2020, the associated event rate of mortality with post-acute COVID-19 sequalae, average time of acquiring/being diagnosed with post-acute COVID-19 sequalae. Secondary objectives included meta-regressing for any possible and predetermined covariates to ascertain of any relationship with post-acute COVID-19 sequalae.

### Quality assessment

Using the NIH Quality Assessment Tool for Observational Cohort and Cross-Sectional Studies^15^, all included publications were reviewed independently for potential risk of bias by two authors (JKM, JMN). The assessment tool uses 14 questions to enable allocation of a score to each article (poor, fair, or good). If there was disagreement regarding the scoring of a study, consensus was met after discussion among both assessors.

### Statistical analysis

Simple descriptive analysis was performed for the five aims of the review. Heterogeneity among the studies was assessed using the chi-squared test and I^2^, however due to suspected variation among the studies and associated heterogeneity random effects models were used for all meta analyses^16^.Post-acute COVID-19 events rates were estimated using random-effects model, the mortality associated with post-acute COVID-19 sequalae was statistically assessed using random effects models (DerSimonian and Laird)^17^, and event rates (ER) were presented. Publication bias was assessed using Begg and Mazumdar Rank Correlation Test and the Egger’s Test of the Intercept and a precision funnel plot was used to ascertain this the publication bias status. To account for any possible heterogeneity (I^2^), sub-group and sensitivity analysis were conducted and in this, some analysis used fixed-effect model analysis, further to these, meta-regression analysis was run for specific pre-determined covariates. For each outcome variable, 95% confidence intervals (CIs) were presented. A p-value < 0.05 was considered statistically significant. The meta-analysis and meta-regression was conducted using Review Manager 5.4 (Cochrane Collaboration, Oxford, UK)^18^ and comprehensive meta-analysis version 4 (CMA v4).

## Results

There were 2197 articles identified in the initial search of databases and reference lists *(Figure 1)*. After initial screening of titles and abstracts 197 articles met the inclusion criteria for review. On full text screening, the number reduced to 57 studies. Further, 14 studies without clinical outcomes were eliminated. A list of the 43 studies ^1,4,27–36,19,37–46,20,47–56,21,57–62,22–26^ that met the inclusion criteria are illustrated in *Table I*.

**Table 1:** Summer of the studies used in the analysis.

|  | *Study/Author* | *Region* | *Study Design* | *Study Setting* | *Average time to PACS diagnosis (in months)* |
| --- | --- | --- | --- | --- | --- |
| 1. | <sup>62</sup> | Africa | Prospective | Single | 2 |
| 2. | <sup>54</sup> | Europe | Prospective | Single | 3 |
| 3. | <sup>55</sup> | Europe | Retrospective | Multicenter | 5 |
| 4. | <sup>51</sup> | USA | Prospective | Multicenter | 2 |
| 5. | <sup>4</sup> | Europe | Prospective | Single | 6 |
| 6. | <sup>43</sup> | Europe | Prospective | Multicenter | 12 |
| 7. | <sup>1</sup> | Asia | Prospective | Single | 6 |
| 8. | <sup>52</sup> | Europe | Prospective | Multicenter | 12 |
| 9. | <sup>50</sup> | Africa | Prospective | Single | 3 |
| 10. | <sup>45</sup> | Asia | Retrospective | Single | 9 |
| 11. | <sup>30</sup> | Europe | Cross sectional | Single | 11 |
| 12. | <sup>19</sup> | Asia | Retrospective | Single | - |
| 13. | <sup>27</sup> | USA | Prospective | Single | 4 |
| 14. | <sup>29</sup> | Europe | Retrospective | Multicenter | 6 |
| 15. | <sup>42</sup> | USA | Retrospective | Multicenter | 1 |
| 16. | <sup>38</sup> | Asia | Prospective | Multicenter | 3 |
| 17. | <sup>37</sup> | Asia | Cross sectional | Multicenter | 3 |
| 18. | <sup>24</sup> | USA | Retrospective | Single | - |
| 19. | <sup>28</sup> | USA | Prospective | Single | 6 |
| 20. | <sup>46</sup> | USA | Cross sectional | Single | - |
| 21. | <sup>39</sup> | Europe | Retrospective | Single | 12 |
| 22. | <sup>36</sup> | Asia | Prospective | Single | - |
| 23. | <sup>35</sup> | Europe | Prospective | Multicenter | 7 |
| 24. | <sup>40</sup> | Europe | Prospective | Multicenter | 3 |
| 25. | <sup>32</sup> | Europe | Prospective | Single | 1 |
| 26. | <sup>33</sup> | Asia | Prospective | Single | - |
| 27. | <sup>48</sup> | Africa | Retrospective | Single | 5 |
| 28. | <sup>56</sup> | Africa | Retrospective | Single | 5 |
| 29. | <sup>60</sup> | Asia | Prospective | Single | - |
| 30. | <sup>34</sup> | Europe | Bidirectional<br>prospective | Single | 6 |
| 31. | <sup>61</sup> | Europe | Cross sectional | Single | 7 |
| 32. | <sup>23</sup> | USA | Retrospective | Multicenter | 5 |
| 33. | <sup>44</sup> | Europe | Prospective | Single | 4 |
| 34. | <sup>25</sup> | Europe | Retrospective | Multicenter | 4 |
| 35. | <sup>26</sup> | Asia | Prospective | Single | 3 |
| 36. | <sup>57</sup> | Europe | Prospective | Multicenter | 3 |
| 37. | <sup>22</sup> | USA | Retrospective | Single | - |
| 38. | <sup>58</sup> | USA | Retrospective | Multicenter | 6 |
| 39. | <sup>47</sup> | Asia | Cross sectional | Single | 4 |
| 40. | <sup>41</sup> | Europe | Prospective | Single | 3 |
| 41. | <sup>21</sup> | USA | Prospective | Multicenter | - |
| 42. | <sup>59</sup> | Europe | Prospective | Single | 3 |
| 43. | <sup>31</sup> | Europe | Retrospective | Single | 9 |

**Figure 1:**
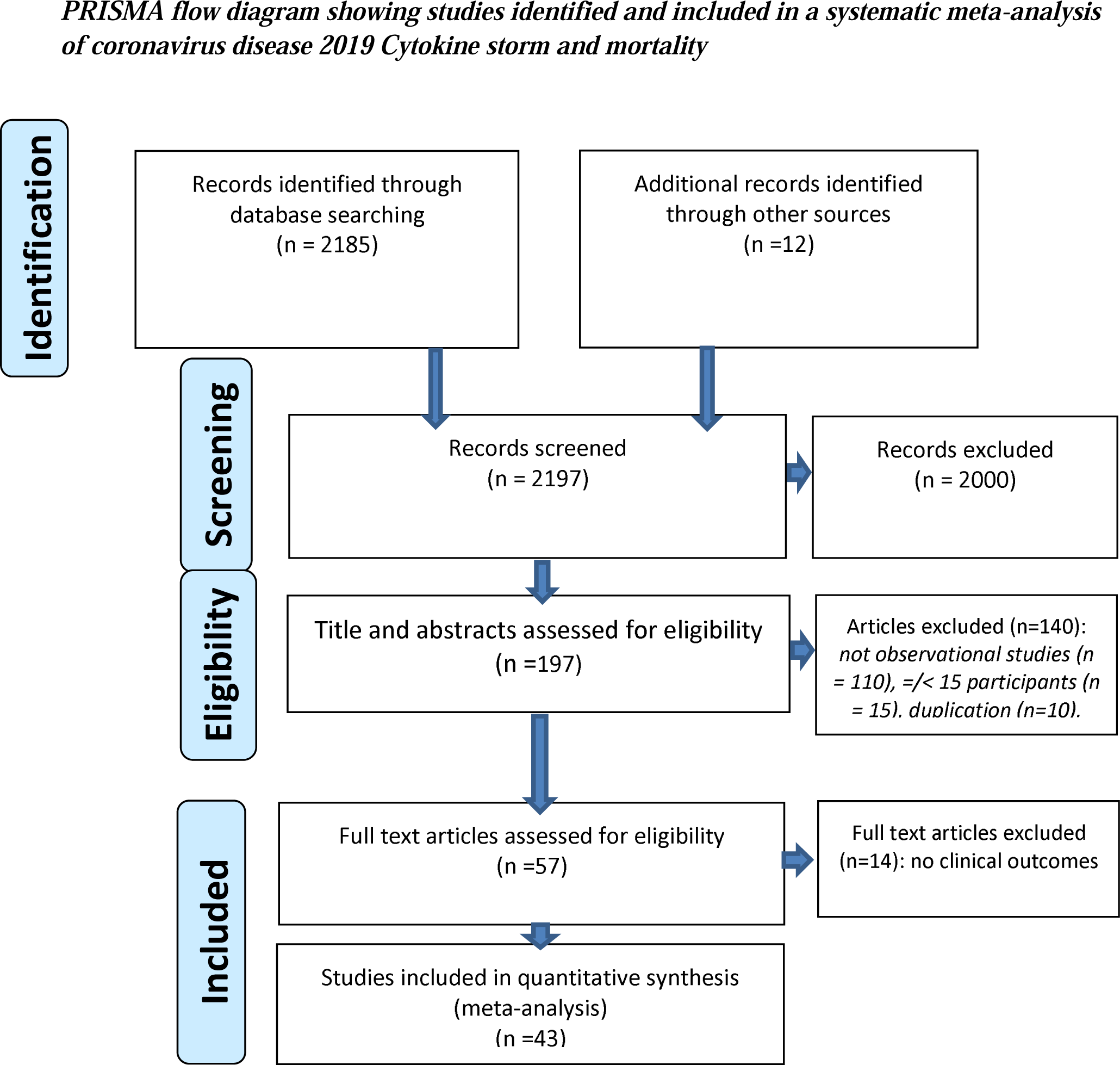
Preferred Reporting Items for Systematic Review and Meta-Analysis (PRISMA) flow diagram for the studies identified and included in the review.

### Prevalence of Post-Acute COVID-19 Sequelae in general population

A total of 43 studies ^1,4,27–36,19,37–46,20,47–56,21,57–62,22–26^ (n= 367236) reported event rates of post-acute COVID-19 Sequelae (n = 183064) in general population. The prevalence of post-acute COVID-19 Sequelae ranged from 1.6% to 82%, with a mean of 42.5% (95% CI 36 % to 49.3%)[Heterogeneity: Tau^2^ = 0.81; Chi^2^ = 24108.789, df = 42 (P = 0.030); I^2^= 100%] *(Figure 2)*. The prediction interval (Figure 3) demonstrates the true effects size in 95% of all the comparable populations fell between 0.10 to 0.823 demonstrating that, in some populations, the event rates of PASC is at one extreme as low as 10% and as high as 82%, a fact that would necessitate a meta-regression analysis to account for possible moderators this variation reflects differences in real proportions.

**Figure 2:**
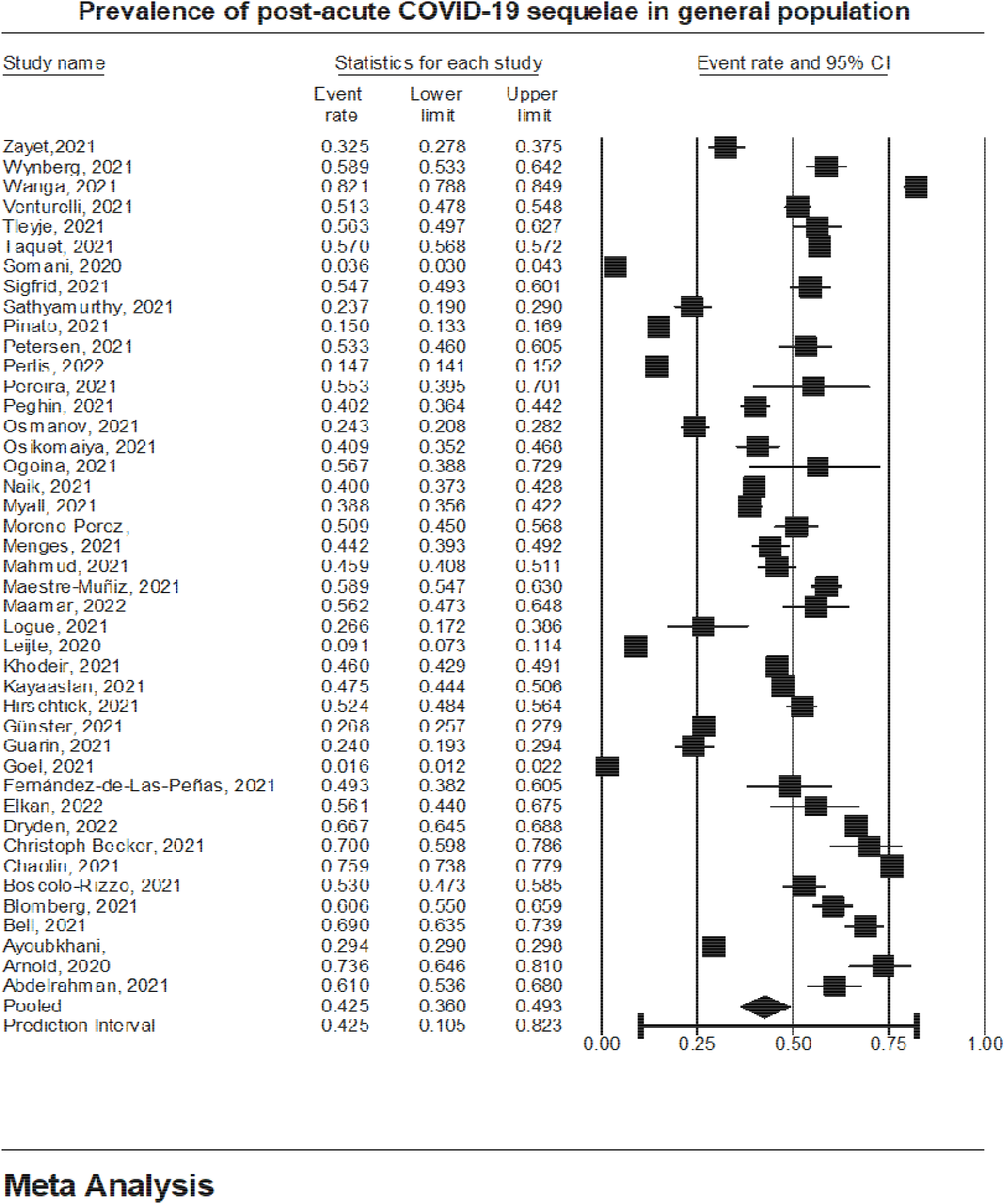
A forest plot on Prevalence of Post-Acute COVID-19 Sequelae in general population

**Figure 3:**
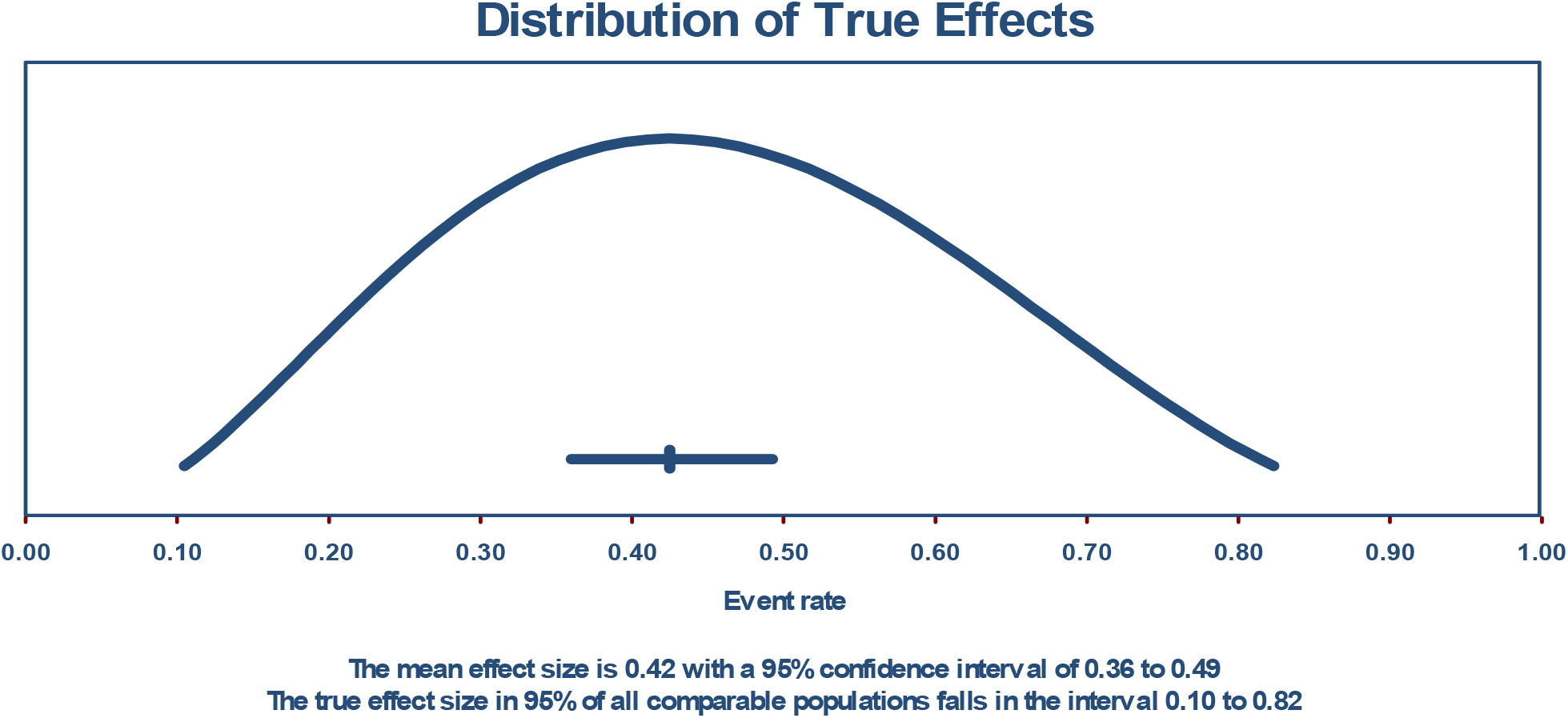
The prediction interval demonstrating the true effects size in 95% of all the comparable populations

A funnel plot of standard error and the egger’s regression intercept test indicated a possible publication bias (intercept = −7.13010, 95% confidence interval (−15.04711, 0.78691), with t=1.81881, df=41 and 1-tailed P = 0.038) *(Figure 4)*.

**Figure 4:**
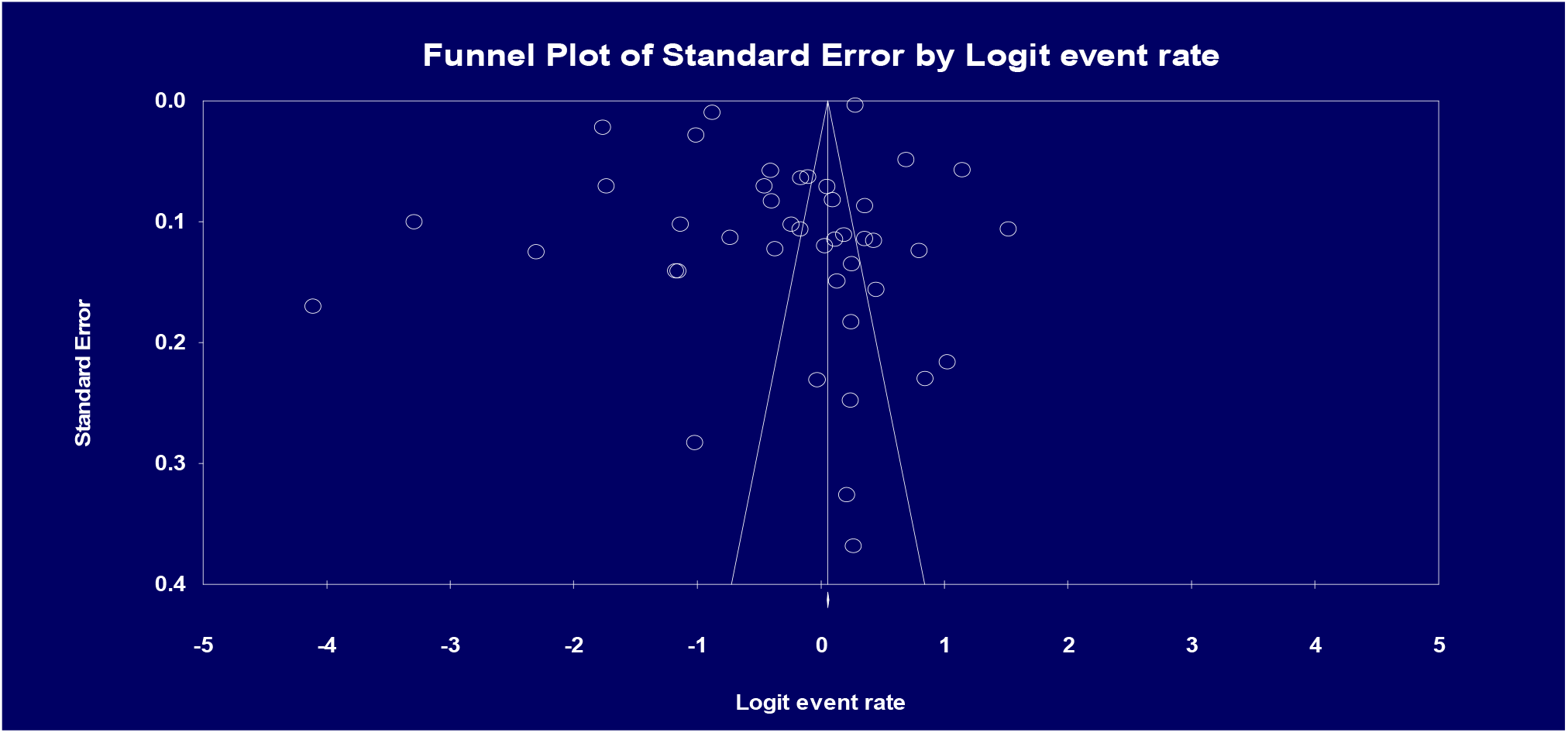
A funnel plot of standard error for publication bias assessment

Sensitivity analysis was performed to explore the impact of excluding or including studies based on sample size, methodological quality and variance on the heterogeneity obtained (I^2^ = 100%). This, by weight allocation for each study using the scale relative to maximum demonstrated that, all contributed between 2% to 3% of the mean event rate and heterogeneity. To further the robustness of this, sensitivity analysis by removing one study with the highest event rate (82%) ^21^,the new mean event rate was 41.4% (95% CI 34.9 %, 448.2%), a non-significant difference from the original 42.5 %. This showed that, the mean PACS event rate estimation was reliable as all the studies depicted a significant P-value *(Figure 5)*.

**Figure 5:**
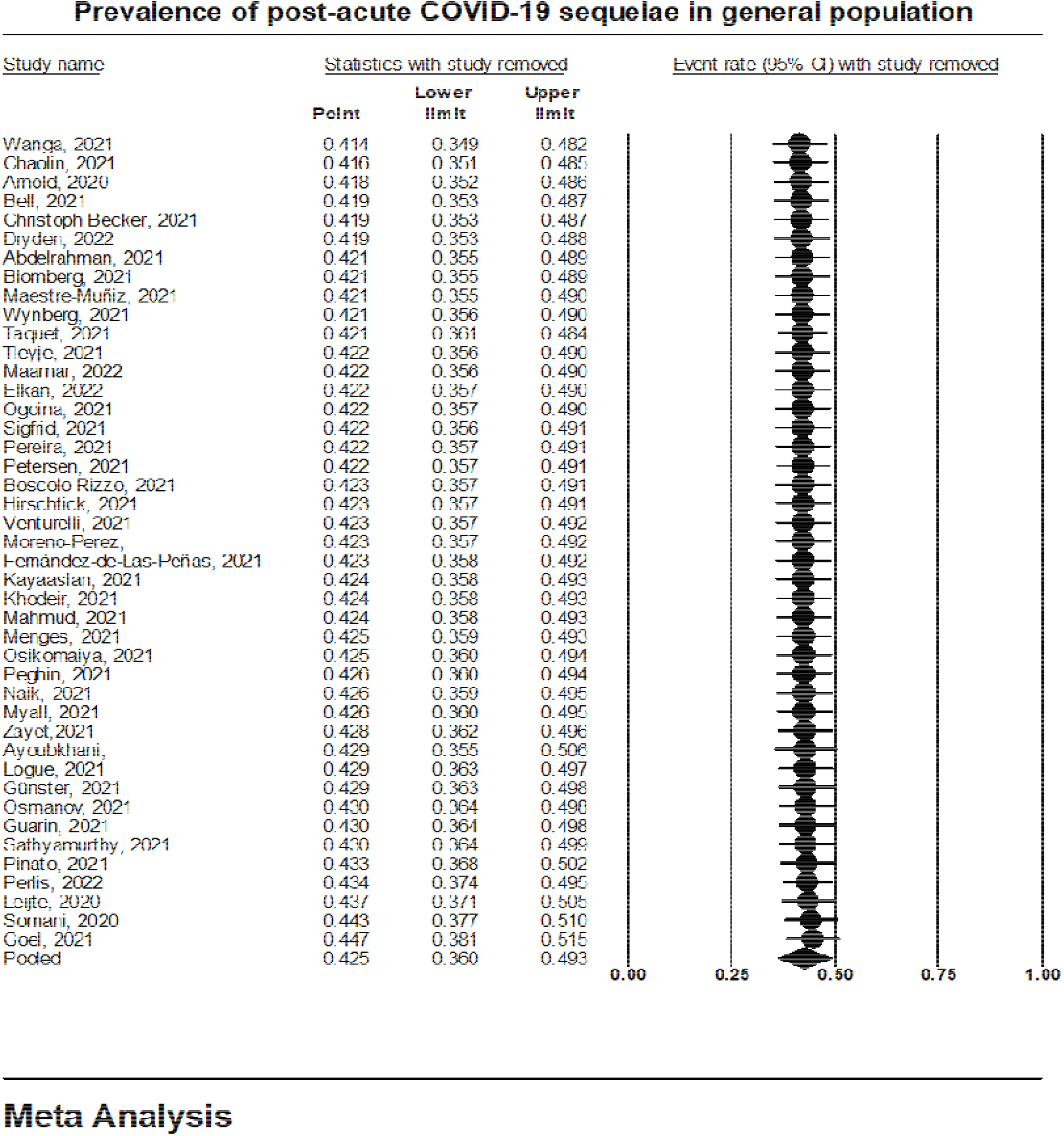
A sensitivity analysis forest plot on prevalence of post-Acute COVID-19 Sequelae in general population

### Average time to and association with post-acute COVID-19 sequalae

Of the 43 studies, thirty-five (35)^1,4,34,35,37–44,23,45,47,48,50–52,54–57,25,58,59,61–66,26–29,31,32^, had demonstrated the times at which either a symptom of PACS was feasible or PACS was diagnosed or some clinical parameters were used to ascertain a case as PACS, while eight^19,21,22,24,33,36,46,60^ did not have any estimated time to PACS diagnosis. Using fixed effect model, the PACS event rates’ range following sub-group analysis was between 25 % (lowest) at four months^25,27,44,47^ and 66 % (highest) at two months^51,62^. Generally, using the point estimates with a fixed effect model, the event rates of PASC was at an average of; one month,44.6%^32,42^, two months, 66%^51,62^, three months, 54.3%^26,37,38,40,41,50,54,57,59^, four months, 25%^25,27,44,47^, five months, 26%^23,48,55,56^, six months, 56.5%^1,4,28,29,34,58^, seven months, 45.1%^35,61^, nine months, 36.3%^31,45^, eleven months,49.3%^63^ and twelve months, 57.9%^39,43,52^.It was evident that, majorly COVID-19 patients developed signs and symptoms of PASC at three months as shown by nine studies (at 54.3%, P= 0.0000) ^26,37,38,40,41,50,54,57,59^ to six months as shown by four studies (at 57%, P= 0.000) ^4,28,34,58^, further up to 12 months^39,43,52^, PACS tends to increase up to (57.9%, P= 0.0148). At an average of two months, however with the highest event rate (66%) in two studies^51,62^, it was not significantly associated with PACS diagnosis (P=0.08) as indicated in the meta-analysis grid (*Table 2*).

**Table 2:** A meta-analysis grid of sub-group analysis on the average time to and association with post-acute COVID-19 sequalae incidence in general population.

|  |  |  |  |  |  |  |  |  |
| --- | --- | --- | --- | --- | --- | --- | --- | --- |
| Fixed effect analysis |  |  |  |  |  |  |  |  |
| Groups by no. of Months |  | Effect size and 95% interval |  |  | Other heterogeneity statistics |  |  |  |
|  | Number of Studies | Point estimate | Lower limit | Upper limit | Q-value | df (Q) | P-value | I-squared |
| - | 8 | 0.28945855 | 0.27537832 | 0.303957059 | 1731.420816 | 7 | 0 | 99.59570776 |
| 1 | 2 | 0.44501583 | 0.4196708 | 0.471630162 | 25.87689388 | 1 | 3.64E-07 | 96.13554855 |
| 11 | 1 | 0.493333333 | 0.38240733 | 0.604919604 | 2.52E-17 | 0 | 1 | 0 |
| 12 | 3 | 0.579451348 | 0.547397643 | 0.610848802 | 8.550605364 | 2 | 1.39E-02 | 76.60984596 |
| 2 | 2 | 0.660100818 | 0.616126929 | 0.701478209 | 3.038312329 | 1 | 7.98E-02 | 67.40879374 |
| 3 | 9 | 0.543060601 | 0.530061253 | 0.556001565 | 272.5184642 | 8 | 0 | 97.06441909 |
| 4 | 4 | 0.25081574 | 0.231004582 | 0.270390787 | 247.8795235 | 3 | 0 | 98.78973464 |
| 5 | 4 | 0.26504428 | 0.261575036 | 0.268542802 | 1356.458021 | 3 | 0 | 99.77883577 |
| 6 | 6 | 0.564949317 | 0.563112141 | 0.566784711 | 2316.437553 | 5 | 0 | 99.78415131 |
| 7 | 2 | 0.451486952 | 0.404547885 | 0.499305419 | 1.701618204 | 1 | 0.192076487 | 41.23241056 |
| 9 | 2 | 0.362924343 | 0.317570851 | 0.410855409 | 12.7830889 | 1 | 3.50E-04 | 92.17713403 |
| Total within |  |  |  |  | 5976.694848 | 32 | 0 |  |
| Total between |  |  |  |  | 18132.09461 | 10 | 0 |  |
| Overall | 43 | 0.514446277 | 0.512793646 | 0.516132579 | 24108.78946 | 42 | 0 | 99.82578968 |

### Sub-group and meta-regression analysis

To account for the substantial unaccounted substantial heterogeneity, it deemed applicable to better understand whether and which study-level factors drove the measures of effect. This meta-regression analysis was on the prevalence of post-acute COVID-19 sequalae in general population and the average time to PACS diagnosis.

On the prevalence of post-Acute COVID-19 sequalae in general population, seventeen studies^4,22,43–46,55,59,60,25–28,31,33,39,42^, detailing a specific leading comorbidity in each study were used for the analysis to determine their association with PACS event rates. On this meta-regression collectively, the comorbidities contributed to 14% of the PACS event rates however, not significant (Q = 7.05, df = 8, p = 0.5313) (R^2^=0.14) *(Supp. File 1)*.

Sub-group analysis on specific comorbidity showed that, a cardiovascular disorder, especially hypertension as per the studies included was significantly associated with post-acute COVID-19 syndrome on contrary to the other conditions depicting that, the proportion of the 14% contributed by the comorbidities was majorly by a patient who had a cardiovascular disease (CVD), majorly hypertension with a stand-alone event rate of 32%, random effects model (0.322 (95% CI 0.166, 0.532) (P = 0.000); I^2^ = 99%). Chronic obstructive pulmonary disorder and abnormal basal metabolic index had higher event rates (59.8 % and 55.9 %), however not significantly associated with PACS (P > 0.05) *(Table 3)*.

**Table 3:** Subgroup meta-analysis grid on comorbidities relative to PACS event.

| Groups | Effect size and 95% interval |  |  | Prediction in Prediction interval |  | Other hetero | Other heterogeneity statistics |
| --- | --- | --- | --- | --- | --- | --- | --- |
| Group | Point estimate | Lower limit | Upper limit | Lower limit | Upper limit | P-value | I-squared |
| **Fixed effect analysis** |  |  |  |  |  |  |  |
| - | 0.548541313 | 0.547743386 | 0.551337947 |  |  | 0 | 99.78881744 |
| Abnormal BMI | 0.558468219 | 0.520029766 | 0.598170102 |  |  | 0.139267521 | 54.25505427 |
| Allergic Conditions | 0.243243243 | 0.208218548 | 0.282060706 |  |  | 1 | 0 |
| Cancer | 0.150289017 | 0.133387586 | 0.168914595 |  |  | 1 | 0 |
| Chronic rhinosinusitis | 0.324858757 | 0.278087973 | 0.375405216 |  |  | 1 | 0 |
| COPD | 0.595296009 | 0.562808546 | 0.627728426 |  |  | 0.637100738 | 0 |
| CVD | 0.280787802 | 0.262533567 | 0.298795302 |  |  | 0 | 99.15520841 |
| Diabetes | 0.294265383 | 0.290195828 | 0.298368018 |  |  | 1 | 0 |
| Hypothyroidism | 0.400324149 | 0.373322562 | 0.427945061 |  |  | 1 | 0 |
| Total within |  |  |  |  |  | 0 |  |
| Total between |  |  |  |  |  | 0 |  |
| Overall | 0.514446277 | 0.512758646 | 0.516132579 |  |  | 0 | 99.82578968 |
| **Random effects analysis** |  |  |  |  |  |  |  |
| - | 0.46739156 | 0.369170412 | 0.568208904 | 9.29E-02 | 0.88261858 |  |  |
| Abnormal BMI | 0.558817678 | 0.228585829 | 0.844631452 | 8.73E-02 | 0.948973318 |  |  |
| Allergic Conditions | 0.243243243 | 3.96E-02 | 0.715003771 | 1.60E-02 | 0.864375156 |  |  |
| Cancer | 0.150289017 | 2.23E-02 | 0.578677645 | 8.87E-03 | 0.77747804 |  |  |
| Chronic rhinosinusitis | 0.324858757 | 5.79E-02 | 0.79009453 | 2.37E-02 | 0.905269402 |  |  |
| COPD | 0.597548617 | 0.257748292 | 0.863919848 | 0.100690644 | 0.951667211 |  |  |
| CVD | 0.322075585 | 0.18579648 | 0.48726487 | 4.82E-02 | 0.81671629 |  |  |
| Diabetes | 0.294265383 | 5.12E-02 | 0.763199677 | 2.07E-02 | 0.881411283 |  |  |
| Hypothyroidism | 0.400324149 | 0.079274327 | 0.838080989 | 0.032724804 | 0.9294401 |  |  |

Hospital re-admission on the other side contributed to 17% of the post-acute COVID-19 syndrome on meta-regression (R^2^ analog = 0.17), and was significantly associated (Q = 8.70, df = 1, p = 0.0032) *(Supp. File 2)*. Study design used in a specific study showed that, the coefficient for retrospective study design (Y) was −0.6815, with a 95% confidence interval of −2.5795 to 1.2165. Studies that used this design had a mean effect size 0.6815 points lower than studies which did not use this design demonstrating that, retrospective study design was probably associated with a smaller effect size on post-acute COVID-19 syndrome. However, there was a significant relationship between the study design and post-acute COVID-19 syndrome (Q = 14.32, df = 3, p = 0.0025) (R^2^=0.26) *(Supp. File 3)*. The study setting whether single site or multiple site did not have any effect on post-COVID-19 syndrome same case for the year a study was conducted.

Collectively, the combined impact of all covariates in the model showed that the model is able to explain at least some of the variance in effect size on post-acute COVID-19 syndrome contributing to 53% of the post-acute COVID-19 syndrome (Q = 38.81, df = 19, p = 0.0047) (R^2^ analog = 0.53) *(Supp. File 4)*.

## Discussion

This review, meta-analysis and meta-regression has demonstrated the prevalence of post-acute COVID-19 sequalae in general population to be 42.5% ranging between1.6% to 82%, and comorbidities contributed to 14% of the PACS event rates specifically, a cardiovascular disorder mostly hypertension which had a stand-alone event rate of 32%. Chronic obstructive pulmonary disorder and abnormal basal metabolic index had higher event rates (59.8 % and 55.9 %), however not significantly associated with PACS. Seventeen percent (17%) of the post-acute COVID-19 sequalae was found to be due to hospital re-admission. Further, there was a significant relationship between the study design and post-acute COVID-19 syndrome while study setting and year a study was conducted did not have any effect on post-COVID-19 syndrome. Generally, other covariates related to either methodological and clinical characteristics were able to explain at least some of the variance in effect size contributing to 53% of the post-acute COVID-19 syndrome.

The reported prevalence of post-acute COVID-19 sequalae in the current review and meta-analysis of 42.5% after index infection with COVID-19 infection was similar to a study that looked at global estimated pooled PASC prevalence derived from the estimates presented in 29 studies as 43%^67^, and primary studies also showed 44.2% reported persistent symptoms post primary infection with COVID-19 disease^68^, as seen in yet another, 49.2% had 3 or more symptoms of COVID-19 post primary infection^69^ and 41% of long COVID-19 with symptoms persisting for more than four weeks^70^. Further, another study, established that, 36.55% of post-acute COVID-19 sequalae occurred between 3 and 6 months and 57.00% had one or more long-COVID feature recorded during the whole 6-month period^58^. Similarly or close to the current findings in this study, another study^71^, established that, among 127 patients who had recovered from COVID-19, 52.0% had persistent symptoms while those with mild COVID-19 recorded 49.5%. Again, conforming to be within the range of post-acute-COVID-19 syndrome in this study (1.6% to 82 %), another study established the prevalence of post-COVID-19 to be 72.6% and 46.2% for hospitalized and non-hospitalized patients, respectively^72^.

On comorbidities commensurate with this current finding, cardiovascular disease (CVD) emerged as a risk factor for the post-acute COVID-19 syndrome, which can affect up to 54% of patients who recover from the acute infection^73^, substantiated further by findings that, presence of certain chronic condition is linked with post-acute COVID-19 sequalae^74^. On the other hand, as found in this study at an event rate of 60 % though not significantly associated with PACS, chronic obstructive pulmonary disorder was found to be among comorbidities associated with long COVID-19 presenting with chronic fatigue (1.68, 1.21 to 2.32)^75^, and again regarding chronic obstructive pulmonary disorder and abnormal basal metabolic index, a study established that, high BMI and previous pulmonary disease could be risk factors for post-acute COVID-19 (long COVID-19) development^76^, further on basal metabolic index, a study concludes that, long COVID is more likely to occur in people with a higher body mass index (BMI)^77^. Like in this study subject to comorbidities generally predicting post-acute COVID-19 sequalae at 14 % (R^2^=0.14), another study established overall severity of comorbidities as one the strongest risk factors for post-acute COVID-19 sequelea^72^. Hospital re-admission contributing to over 17 % of post-acute COVID-19 sequalae as per this study, similar findings have shown some consistency where patients (19.9%) who survived COVID-19 hospitalization were readmitted^78^, and another, the readmission rate was 13.3%^79^.

This study reported COVID-19 patients developed signs and symptoms post-acute COVID-19 syndrome at three months (at 54.3%) to six months (at 57%) which tended to increase up to 12 months (57.9%). Similar or close to these findings, another study established that, at 3 months of follow-up, 66·7%) of participants reported new or persistent COVID-19-related symptoms^50^ and another that, hospitalized and non-hospitalized patients with confirmed or suspected COVID-19, multiple symptoms are present about 3 months after symptoms onset^80^. A forty-five per cent (45%) of health care workers also reported persistent symptoms at three to four months^81^. As regards post COVID-19 sequalae at six months (6-months), a previous study shows post-acute COVID-19 sequalae prevalence was 50% in adults at 6 months^82^, close to the 54% in the current findings. At 12 months, post-acute COVID-19 sequalae was reported as (50%) ≥1 PASC symptom, close to 58% as per the current findings. This is further substantiated by the findings in a study that, Persistent symptoms were highly prevalent, especially fatigue, shortness of breath, headache, brain fog/confusion, and altered taste/smell, which persisted beyond 1 year (12 months) among 56%^83^. Contrary to the current findings though, some studies have reported a decreasing post-acute COVID-19 sequalae from sixth to twelfth month ^1,82^

A limitation of the current review was the defined inclusion criteria for a post-acute COVID-19 sequalae patient, presenting with either one or more suspected signs and symptoms depicting the syndrome. This may have resulted in diverse estimate of the prevalence of post – acute COVID-19 during different times to event which were also estimated on an average basis. However, the majority of studies in the current review included a clearly mentioned post-acute COVID-19 sequalae either as persistent symptoms after primary infection with COVID-19 or a cluster of signs and symptoms post recovery from the index infection with COVID-19. Further, the review focused only on the post-acute COVID-19 sequalae devoid categorizing it as per the time a patient experienced the same. This, was even though not very important as the authors intended to ascertain the prevalence and any association with subject variable, plus, a meta-regression was performed to ensure such differential aspects were taken care of.

## Data Availability

All data produced in the present work are contained in the manuscript

## Author contributions

**J.K.M:** Designed the study, reviewed and analyzed the data, participated in quality assessment process, Wrote and submitted the manuscript.

**C.M.M:** Reviewed studies and participated in quality assessment process.

**J.N.M:** Reviewed and analyzed the data, Edited the manuscript.

**J.K.O:** Participated in quality assessment process, Edited the manuscript.

**R.M.N:** Reviewed the studies, participated in quality assessment process, Proofread the manuscript

**M.K.K:** Participated in quality assessment, Edited the manuscript.

## Ethical review statement

No ethical approval was required for this study.

## Study’s Supplementary Files

**Supp. File 1.**
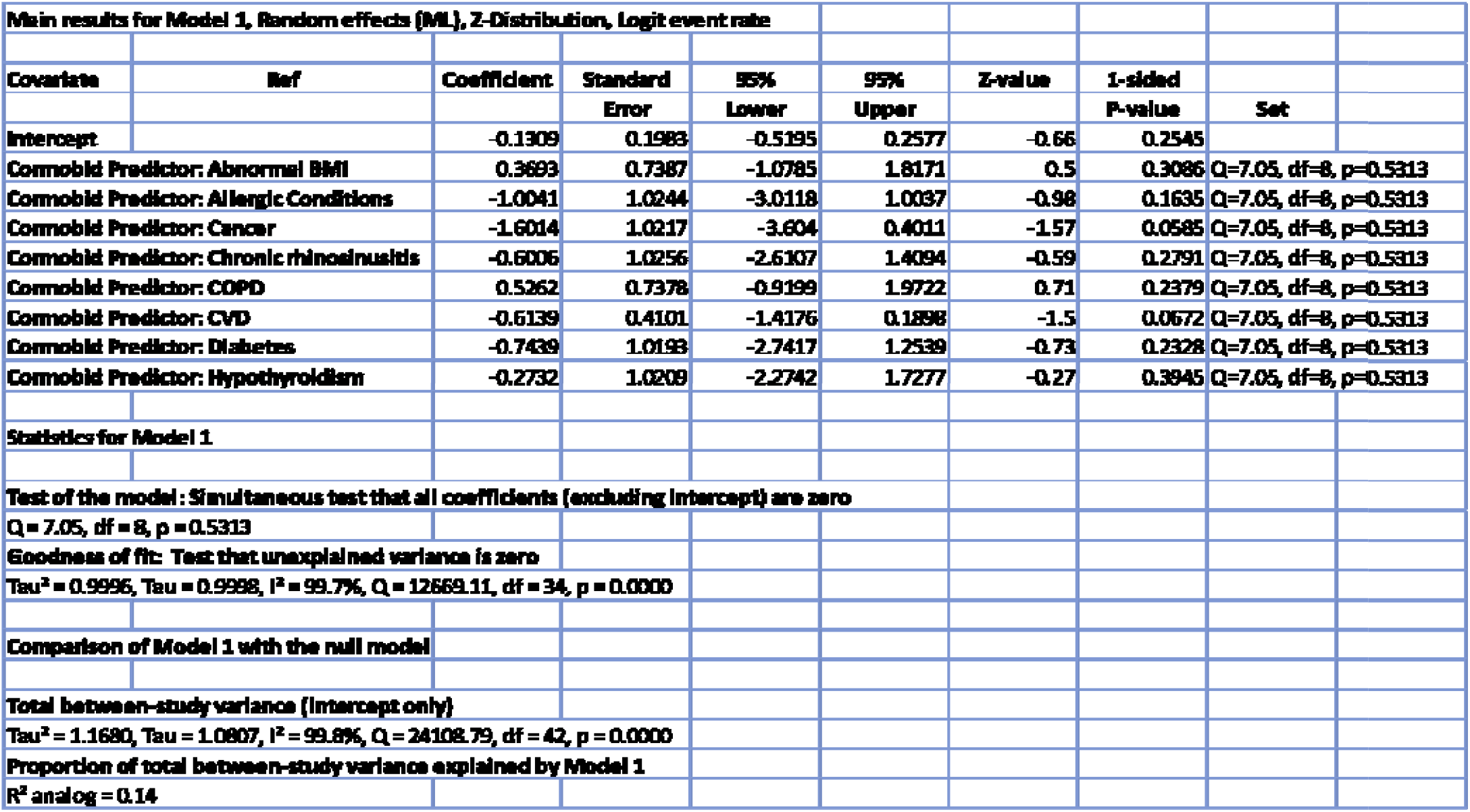

**Supp. File 2.**

| Main results for Model 1, Random effects (ML), Z-Distribution, Logit event rate |  |  |  |  |  |  |  |  |
| --- | --- | --- | --- | --- | --- | --- | --- | --- |
| Covariate | Ref | Coefficient | Standard Error | 95% Lower | 95% Upper | Z-value | 1-sided P-value | Set |
| Intercept |  | -0.1215 | 0.1633 | -0.4416 | 0.1986 | -0.74 | 0.2285 |  |
| Hospital re-admission: Yes |  | -1.2871 | 0.4364 | -2.1425 | -0.4317 | -2.95 | 0.0016 |  |
| Statistics for Model 1 |  |  |  |  |  |  |  |  |
| Test of the model: Simultaneous test that all coefficients (excluding intercept) are zero |  |  |  |  |  |  |  |  |
| Q = 8.70, df = 1, p = 0.0032 |  |  |  |  |  |  |  |  |
| Goodness of fit: Test that unexplained variance is zero |  |  |  |  |  |  |  |  |
| Tau<sup>2</sup> = 0.9652, Tau = 0.9825, I<sup>2</sup> = 99.6%, Q = 11597.69, df = 41, p = 0.0000 |  |  |  |  |  |  |  |  |
| Comparison of Model 1 with the null model |  |  |  |  |  |  |  |  |
| Total between-study variance (intercept only) |  |  |  |  |  |  |  |  |
| Tau<sup>2</sup> = 1.1680, Tau = 1.0807, I<sup>2</sup> = 99.8%, Q = 24108.79, df = 42, p = 0.0000 |  |  |  |  |  |  |  |  |
| Proportion of total between-study variance explained by Model 1 |  |  |  |  |  |  |  |  |
| R<sup>2</sup> analog = 0.17 |  |  |  |  |  |  |  |  |

**Supp. File 3.**

| Main results for Model 1, Random effects (ML), Z-Distribution, Logit event rate |  |  |  |  |  |  |  |
| --- | --- | --- | --- | --- | --- | --- | --- |
| Covariate | Coefficient | Standard Error | 95% Lower | 95% Upper | Z-value | 1-sided P-value | Set |
| Intercept | -0.3957 | 0.9351 | -2.2284 | 1.437 | -0.42 | 0.3361 |  |
| Study design: Cross sectional | 0.4985 | 1.0277 | -1.5157 | 2.5128 | 0.49 | 0.3138 | Q=14.32, df=3, p=0.0025 |
| Study design: Prospective | 0.4826 | 0.9554 | -1.39 | 2.3552 | 0.51 | 0.3067 | Q=14.32, df=3, p=0.0025 |
| Study design: Retrospective | -0.6815 | 0.9684 | -2.5795 | 1.2165 | -0.7 | 0.2408 | Q=14.32, df=3, p=0.0025 |
| Statistics for Model 1 |  |  |  |  |  |  |  |
| Test of the model: Simultaneous test that all coefficients (excluding intercept) are zero |  |  |  |  |  |  |  |
| Q = 14.32, df = 3, p = 0.0025 |  |  |  |  |  |  |  |
| Goodness of fit: Test that unexplained variance is zero |  |  |  |  |  |  |  |
| Tau<sup>2</sup> = 0.8674, Tau = 0.9313, I<sup>2</sup> = 99.8%, Q = 24038.05, df = 39, p = 0.0000 |  |  |  |  |  |  |  |
| Comparison of Model 1 with the null model |  |  |  |  |  |  |  |
| Total between-study variance (intercept only) |  |  |  |  |  |  |  |
| Tau<sup>2</sup> = 1.1680, Tau = 1.0807, I<sup>2</sup> = 99.8%, Q = 24108.79, df = 42, p = 0.0000 |  |  |  |  |  |  |  |
| Proportion of total between-study variance explained by Model 1 |  |  |  |  |  |  |  |
| R<sup>2</sup> analog = 0.26 |  |  |  |  |  |  |  |

**Supp. File 4.**
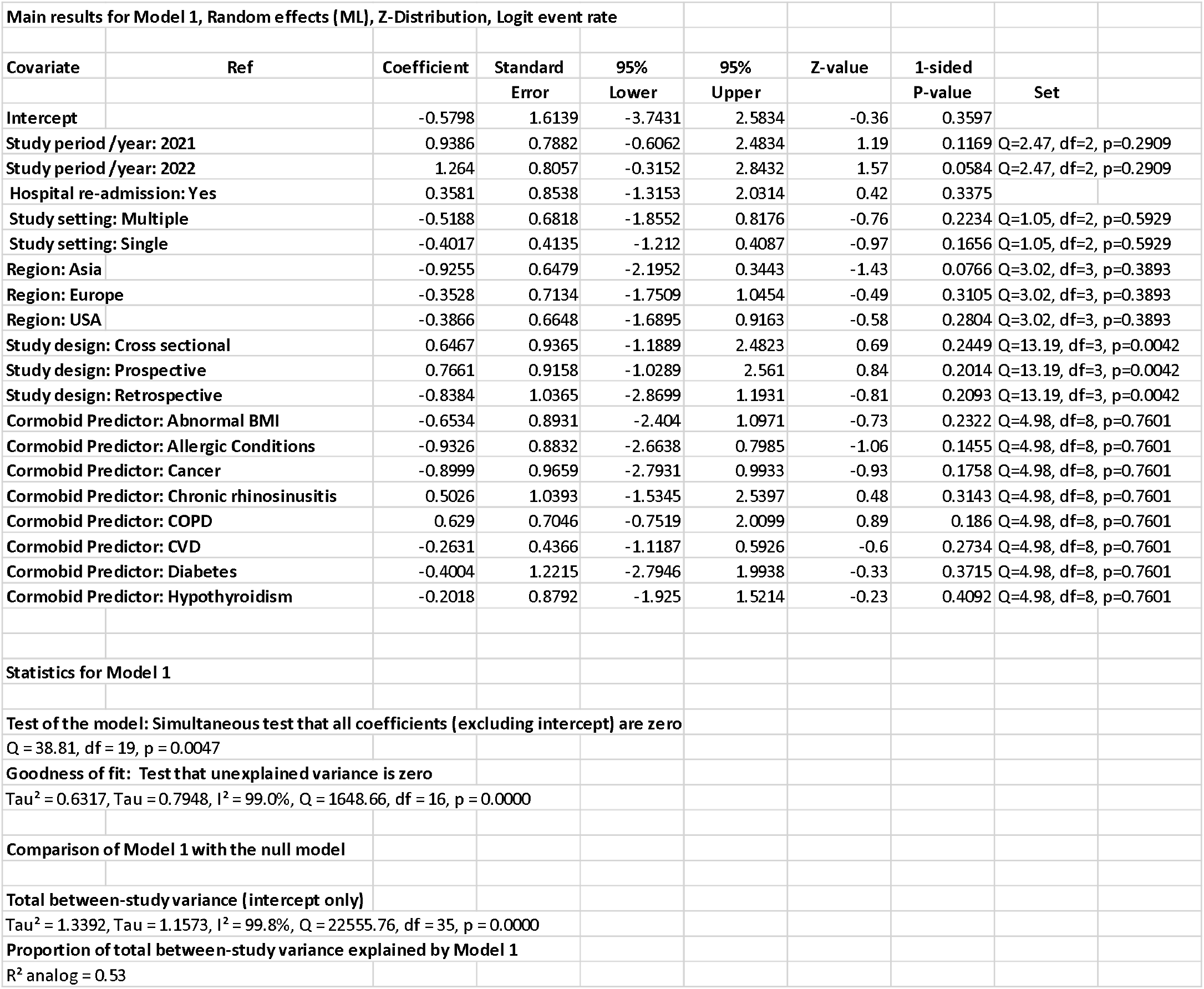

